# Regional Variation in COVID-19 Scarce Resource Allocation Protocols

**DOI:** 10.1101/2021.01.14.21249845

**Authors:** Rupali Gandhi, Gina M. Piscitello, William F. Parker, Kelly Michelson

## Abstract

**Background:** Scarce resource allocation policies vary across the United States. Little is known about regional variation in hospital-level resource allocation protocols and variation in their application.

**Objective:** To evaluate how scarce resource allocation policies throughout the Chicagoland area vary and whether there are differences in policy application within and amongst hospitals.

**Design:** Two cross-sectional surveys.

**Setting:** Chicagoland hospitals.

**Participants:** Representatives from Chicagoland hospitals and triage officers at these hospitals.

**Measurements:** Survey responses and categorical variables are described by frequency of occurrence. Intra- and interhospital variation in ranking of hypothetical patients was assessed using Kappa coefficients.

**Results:** Eight Chicago area hospitals responded to the survey assessing scarce resource allocation protocols. For hospitals willing to describe their specific ventilator allocation protocol (n=7), the initial scoring system varied with most utilizing the sequential organ failure assessment (SOFA) score (86%) and medical comorbidities (57%). A majority gave priority to pre-defined groups in their initial scoring system (86%), all discussed withdrawal of mechanical ventilation for adult patients (100%), and a minority had exclusion criteria (43%). Forty-nine triage officers from nine hospitals responded to the second survey. Triage officer rankings of hypothetical patients had slight agreement amongst all hospitals (Kappa 0.158) and fair agreement within two hospitals with the most respondents (Kappa 0.21 and 0.25). Almost half of triage officer respondents reported using tiebreakers to rank patients (N=23/49, 47%).

**Conclusion:** Although most Chicago area hospitals surveyed created guidelines for resource allocation during the COVID-19 pandemic, these guidelines and application of these protocols by triage officers varied significantly.

**Funding Source:** None.

## Introduction

Concern about scarcity of resources such as mechanical ventilators during the Coronavirus Disease 2019 (COVID-19) pandemic required United States (US) hospitals to create resource allocation policies in preparation for critical care shortages. Experts proposed multiple ideas for equitably allocating ventilators^1–8^ without consensus. US State resource allocation policies vary widely,^9,10^ but the range in these protocols within states or individual cities is not well known. In addition, little is known about the reliability in implementation of these protocols by those asked to utilize them.

Like other metropolitan areas, Chicago has many hospitals ranging from community hospitals to large tertiary care centers.^11^ Often patients receive care at more than one hospital. Large variation in resource allocation – especially critical care resources – amongst hospitals in the same city, could have important consequences. There could be concerns about inequitable treatment of certain populations.^12^ Furthermore, the principled reasoning behind creating resource allocation policies could be rendered obsolete because patients could choose to go a hospital where they knew they would get priority.

This study aims to evaluate the practical effect of resource allocation policies at hospitals in the Chicago metropolitan area, including initial scoring systems, exclusion criteria, priority groups, and tiebreakers by using a cross sectional survey of institutional representatives at these hospitals. It also assesses how different patients would be prioritized when using these protocols and evaluates if differences in allocation might occur within and across hospitals by surveying potential triage officers, people assigned to determine allocation priority, at Chicago area hospitals.

## Methods

A cross-sectional survey study was used to evaluate scarce resource allocation protocols at hospitals or hospital systems in and near Chicago, Illinois. The primary aim was to describe and contrast ventilator allocation policies throughout the Chicagoland area. The study also evaluated how patients would be triaged using these policies in practice and assessed if differences in allocation occurred within and amongst hospitals.

### Recruitment of Participants and Survey Design

Two surveys were designed using best practice after completing a literature review.^13^ These surveys were primarily designed by a group of physicians including a palliative medicine physician (GMP), a pediatric cardiologist (RG) and a pulmonary critical care physician (WFP) all of whom have experience in ethics and contributed to developing their own hospital resource allocation protocols. Institutional review board (IRB) exemption was granted by Lurie Children’s Hospital of Chicago IRB.

Surveys were sent by email during August and September 2020 using REDCap hosted by Northwestern University.^14,15^ The first survey aimed to evaluate specific details of ventilator and other scarce resource allocation policies. This survey included 53 multiple choice and free-response questions on the initial scoring system, exclusion criteria, priority groups, and tiebreakers of allocation protocols. For each hospital, this survey was sent to a physician or ethicist who had access to their respective institution’s ventilator allocation protocol. Surveys did not request information that would identify the source hospital, healthcare system, or respondent.

The second survey evaluated triage officers’ execution of their institution’s resource allocation protocol. The survey included 17 multiple choice and free-response questions including a question asking the respondent to rank 6 hypothetical patients from highest to lowest priority for receiving scarce resources using their own institution’s allocation policy. For each hypothetical patient, age, job description, brief past medical history and current sequential organ failure assessment (SOFA)^16^ score were provided (**Table 1**). A higher SOFA score correlates with higher likelihood of mortality.^16^ Age and job description were included because some hospital protocols were thought to be likely to use these when determining priority scores or as tiebreakers if two patients were of equal priority. For the hypothetical pediatric patient, several different pediatric prognostic scores found in the literature^17–21^ were provided along with a mortality estimate since there is more variability among pediatric protocols. ^9^ The authors identified one person at Chicago-area institutions where a resource allocation protocol was known to exist and asked that person to distribute the survey to people that the hospital identified to participate in triaging patients (i.e. triage officers). This method was used to maximize the response rate among triage officers as it allowed the respondents to remain completely anonymous to the authors and no identifying information about respondents was provided to the authors.

**Table 1:**
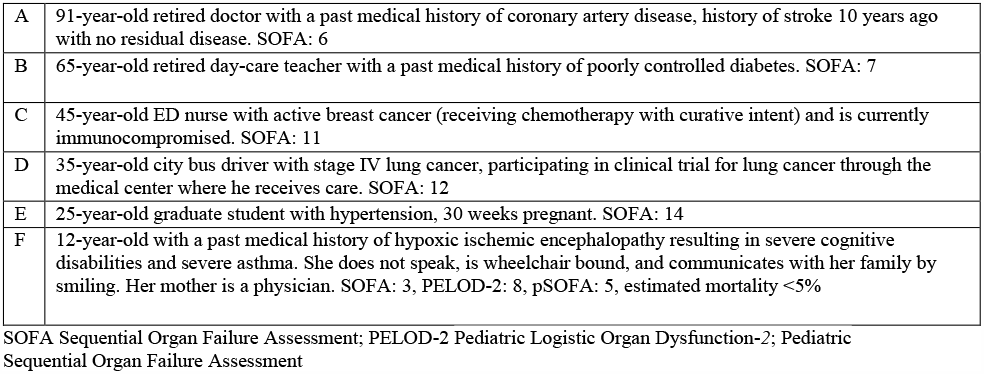
Six Hypothetical Patients

### Statistical Analysis

Survey responses and categorical variables are described by frequency of occurrence. For each hypothetical patient, the median, mean, and modal rank were determined. Because each rater scored 6 patients on an ordinal scale, Fleiss’s kappa coefficients were calculated to assess for agreement among respondents.^22^ A kappa of less than or equal to zero indicates no agreement (other than what would be expected by chance) and a kappa of one indicates complete agreement. All analyses were conducted using R version 3.6.1 (The R Foundation for Statistical Computing, Vienna, Austria). All statistical testing was 2-sided with a *P*-value threshold of < 0.05.

## Results

### Hospital Characteristics

Eight Chicago area hospitals or hospital systems responded to the first survey (N=8/18, response rate 44%). Nine Chicago-area hospitals had triage officers who responded to the second survey (N=9/10, response rate 90%). The majority of respondent hospital or hospital systems were private (75%) and academic (100%), and they all trained medical students, residents or fellows (100%). Half had a religious affiliation, and 88% offered clinical trials related to COVID-19 to patients (e-Table 1).

### Survey 1: Ventilator Allocation Protocols

Most respondents (N=7/8, 88%) reported the creation of a ventilator allocation protocol at their hospital or hospital system. For hospitals with allocation policies, the initial scoring system used varied, with most utilizing the SOFA score (N=6/7, 86%) and one using the modified sequential organ failure assessment score (N=1/7, 14%). Most also utilized medical comorbidities in the initial scoring system (N=4/7, 57%) (**Table 2**). A minority of protocols had exclusion criteria (N=3/7, 43%) with specific exclusion criteria varying by hospital (e-Table 2). These exclusion criteria remove patients with certain diagnoses from consideration of mechanical ventilation, such as patients with persistent coma or vegetative states or severe burns with less than 10% chance of survival. Most protocols included priority groups in their initial scoring system (N=6/7, 86%), with a majority giving priority for pregnant patients (N=6/7, 86%) and healthcare workers (N=5/7, 71%), and a minority giving priority to other essential workers (N=2/7, 29%) or families of essential workers (N=1/7, 14%) (e-Table 3-4). All discussed withdrawal of mechanical ventilation for adult patients (N=7/7, 100%). At these hospitals, ventilator withdrawal would be considered if a shortage occurred and a new patient came to the hospital with higher likelihood of recovery than a patient who was currently intubated. The method to withdraw ventilators varied by hospital (e-Table 5). Half of the protocols had unique criteria to triage pediatric patients (e-Table 6), with one hospital giving pediatric patients priority over all other patients (e-Table 4). Tiebreakers varied greatly amongst hospitals with some hospitals giving priority to certain groups in a tie, such as younger patients (N=3/7, 43%), patients who were COVID-19 research subjects (N=1/7, 14%), or their own hospital system front line workers (N=1/7, 14%), and most using random allocation as the final tiebreaker (N=4/7, 57%) (e-Table 7).

**Table 2:**
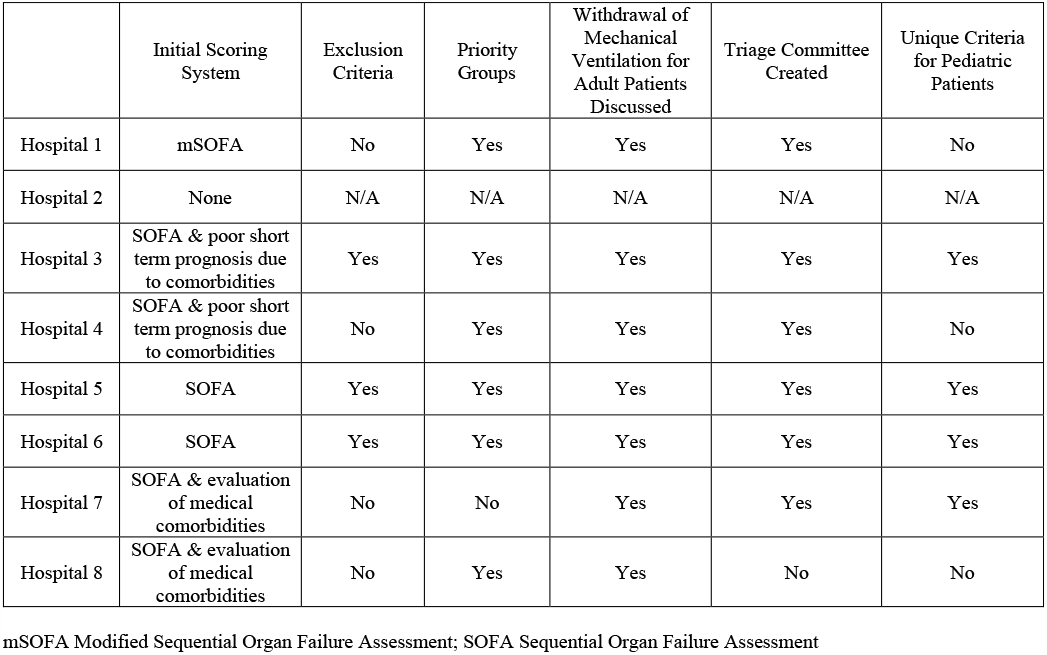
Ventilator Allocation Protocol

|  | Initial Scoring System | Exclusion Criteria | Priority Groups | Withdrawal of Mechanical Ventilation for Adult Patients Discussed | Triage Committee Created | Unique Criteria for Pediatric Patients |
| --- | --- | --- | --- | --- | --- | --- |
| Hospital 1 | mSOFA | No | Yes | Yes | Yes | No |
| Hospital 2 | None | N/A | N/A | N/A | N/A | N/A |
| Hospital 3 | SOFA & poor short term prognosis due to comorbidities | Yes | Yes | Yes | Yes | Yes |
| Hospital 4 | SOFA & poor short term prognosis due to comorbidities | No | Yes | Yes | Yes | No |
| Hospital 5 | SOFA | Yes | Yes | Yes | Yes | Yes |
| Hospital 6 | SOFA | Yes | Yes | Yes | Yes | Yes |
| Hospital 7 | SOFA & evaluation of medical comorbidities | No | No | Yes | Yes | Yes |
| Hospital 8 | SOFA & evaluation of medical comorbidities | No | Yes | Yes | No | No |

mSOFA Modified Sequential Organ Failure Assessment; SOFA Sequential Organ Failure Assessment

### Creation of Ventilator Allocation Protocol

Protocols were created by multidisciplinary groups at each hospital (e-Table 8). Only two hospitals included community input in formation of the protocol (N=2/7, 29%) and a minority had a plan in place to discuss their approach to allocate ventilators with the public (N=3/7, 43%). No institution’s protocol was publicly available (e-Table 9). Only two hospitals rehearsed use of their protocol, e.g., doing trial runs with hypothetical patients (N=2/7, 29%), and no hospital had used their protocol in practice (e-Table 8). Almost all protocols created a triage committee to allocate ventilators (N=6/7, 86%) whose composition varied by hospital (e-Table 10). Information provided to triage committees varied by hospital with some hospitals blinding certain information such as patient name, race and ethnicity (N=2/7, 29%) (e-table 11). In addition to ventilator allocation policies, most hospitals (N=6/8, 75%) created other scarce resource policies (e-table 12). Some of these policies were actually used in practice, such as allocation strategies for Remdesivir and extracorporeal membrane oxygenation.

### Survey 2: Variation in Allocation Amongst Hospital Protocols

For the second survey, there were 49 respondents from 9 Chicago-area hospitals. One survey was removed from analysis because the respondent indicated that she/he had ranked the hypothetical patients at random. The largest number of responses from any one hospital was 20. Twenty-nine respondents were physicians (59%), 8 were nurses or advanced practice nurses (16%), 4 were hospital administrators (8%), and 8 identified as ethicists (16%). Almost all respondents reported being familiar or very familiar with their hospital resource allocation protocol (N=43/49, 88%) and that their institution’s policy provided sufficient guidance to rank the given hypothetical patients (N=43/49, 88%). Few respondents thought that their institution’s policy disadvantaged any group (N=5/49, 10%) and listed racial minorities, low socioeconomic status, elderly age, people with comorbidities, people with disabilities, and women as the groups affected. Most respondents agreed with their institution’s policy to give priority to specific groups such as healthcare workers, pregnant patients or pediatric patients (N=38/49, 78%) and only a minority stated that their institution did not give priority to specific groups (N=10/49, 20%). Only 1 respondent disagreed with how his/her institution gave priority to certain groups.

### Ranking of Six Theoretical Patients

There was pronounced variation amongst respondents’ ranking of the six hypothetical patients. Patient A, a 91-year-old, retired doctor with coronary artery disease and history of a stroke 10 years ago, and SOFA score of 6 was most frequently ranked second (31%). Patient B, a 65-year-old retired daycare teacher with poor controlled diabetes and SOFA 7 was most frequently ranked second and third (29%). Patient C, a 45-year-old ED nurse with active breast cancer, receiving chemotherapy and immunocompromised with SOFA 11 was most often ranked third (35%). Patient D, a 35-year-old, city bus driver with stage IV lung cancer, participating in a clinic trial with SOFA 12 was most often ranked last (49%). Patient E, a 25-year-old graduate student with hypertension and 30-weeks pregnant with SOFA 14 was most often ranked fourth (24%). Finally, patient F, a 12-year-old with hypoxic ischemic encephalopathy, severe cognitive disabilities, who communicates only by smiling, with asthma and estimated mortality <5%, was most often ranked first (43%). The mean and median rank for each patient is reported in **Table 3**.

**Table 3:** Mean and Median Rank for Each Hypothetical Patient with and without Pediatric Patient F

| Patient | With Pediatric Patient Included | Without Pediatric Patient |
| --- | --- | --- |

|  | Mean | Median | Mean | Median |
| --- | --- | --- | --- | --- |
| A | 2.84 | 3 | 2.35 | 2 |
| B | 2.43 | 2 | 1.9 | 2 |
| C | 3.41 | 3 | 2.88 | 3 |
| D | 5.37 | 5 | 4.67 | 5 |
| E | 3.35 | 3 | 2.78 | 3 |
| F | 3 | 2 | N/A | N/A |

Patient F, the pediatric patient, was excluded from entering the allocation protocol according to some respondents due to the patient’s described encephalopathy and therefore, given *lowest* priority. Other respondents excluded this patient from their allocation protocol because she was a pediatric patient and therefore gave this patient the *highest* priority. Figure e-1 shows the distribution of ranking for the pediatric patient with most respondents ranking this patient either 1 or 6. After seeing this wide variation in ranking the pediatric patient, a post-hoc sub analysis was done excluding patient F. When patient F was removed, and the other patients re-ordered as though F had not been a choice, there was slightly less variability (**Figure 1**). Agreement among respondents using Kappa was 0.095 when the pediatric patient was included, and 0.158 when the pediatric patient was excluded (0 = completely random, 1 = fully in agreement) consistent with slight agreement among respondents. Kappa for each rank position when the pediatric patient was excluded was: rank 1 = 0.078, rank 2 = 0.156, rank 3 = 0.090, rank 4 = 0.018, and rank 5 = 0.470 showing that the most agreement was for rank 5 as patient D was most consistently ranked fifth.

**Figure 1:**
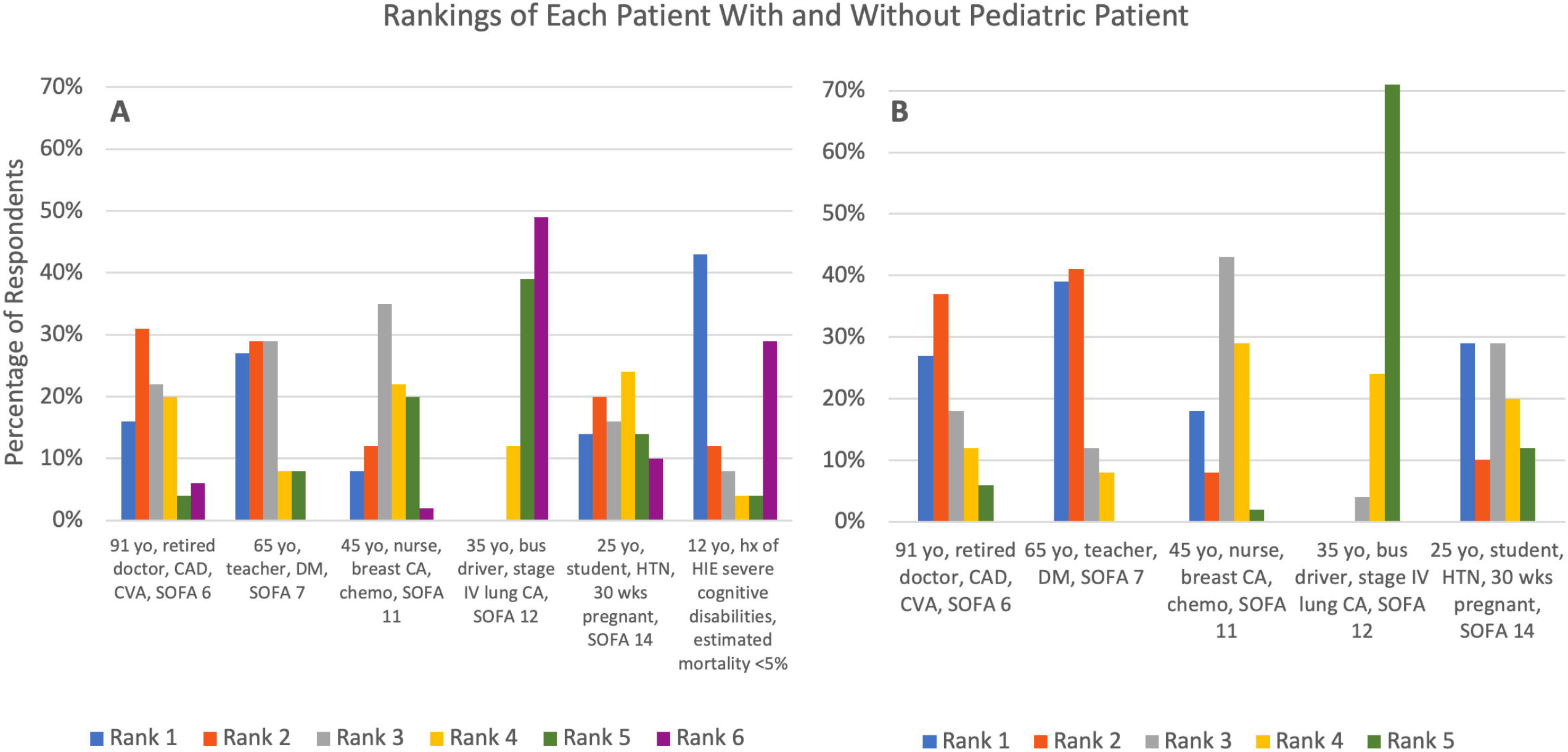
There is great variation in how all respondents ranked the hypothetical patients. (yo = year old, CAD = coronary artery disease, CVA = cerebral vascular accident, SOFA = sequential organ failure assessment, CA = cancer, chemo = chemotherapy, HTN = hypertension, wks = weeks, HIE = hypoxic ischemic injury.)

The variation in ranking existed regardless of whether respondents were from different hospitals or from the same hospital. In a sub-analysis, data from the 2 hospitals with the most respondents (N=20 and N=12) demonstrated that even within the same hospital, there was little agreement amongst respondents (**Figure 2**). Kappa was 0.21 and 0.25 respectively, consistent with only fair agreement amongst respondents.

**Figure 2:**
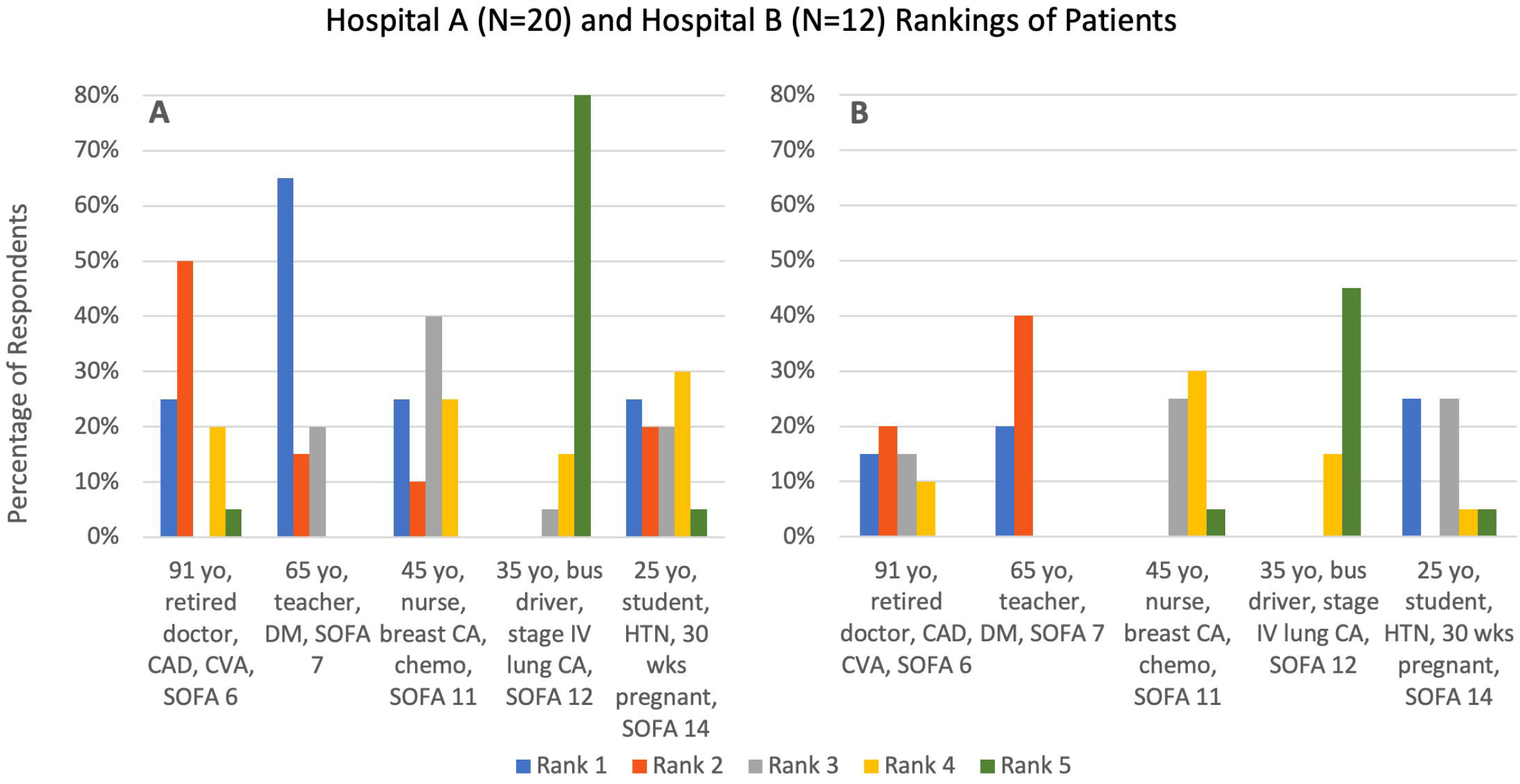
Even within hospitals A and B the variation in ranking persisted. (yo = year old, CAD = coronary artery disease, CVA = cerebral vascular accident, SOFA = sequential organ failure assessment, CA = cancer, chemo = chemotherapy, HTN = hypertension, wks = weeks.)

Almost half of the respondents reported using tiebreakers to assist with the rankings (N=23/49, 47%) and listed using tiebreakers most often between patients C and E (N=7/23, 30%) and patients A and B (N=6/23, 26%). Ties between three or more patients were identified 22% of the time. Respondents used the following criteria to break the ties: age, greatest likelihood of positive outcome, reciprocity, healthcare worker, pregnancy and lottery.

## Discussion

Ventilator allocation policies varied widely amongst hospitals in and surrounding Chicago, Illinois. Although most hospitals used SOFA in their initial scoring system, they differed in that only half also utilized assessment of medical comorbidities in their scoring system. Most protocols gave priority to certain groups such as pregnant patients or healthcare providers. Few protocols were created with community input, and no protocol was available for public review. There was also significant variation in the implementation of resource allocation protocols. There was only slight agreement on the ranking of hypothetical patients among all triage officers, and within two hospitals there was only fair agreement among triage officers. Almost half of triage officer respondents reported using tiebreakers to assist with the rankings.

Although the Illinois Department of Public Health (IDPH) Guidelines on Hospital Emergency Preparedness During COVID-19 make recommendations for how scarce resources should be allocated, the survey data revealed that Illinois hospital protocols are not uniform with these recommendations. The IDPH recommended hospitals make crisis standards of care processes transparent, however no hospital in this study allowed their ventilator allocation policies to be available for public review.^23^ The IDPH guidelines also recommend equity in allocation that does not discriminate based on race, ethnicity, and socioeconomic status, however the majority of ventilator allocation policies utilized medical comorbidities which may lower priority. No protocols introduced efforts to reduce the impact of baseline structural inequities as some experts and ethicists now support.^24^ The IDPH guidelines recommend creation of a triage team consisting of an infectious disease physician, nursing staff, hospital administration and medical ethicists. Survey results showed that although 86% of hospitals created a triage team, only 29% of policies included the members recommended by IDPH.

Most hospitals surveyed gave priority to certain groups in the population over others for mechanical ventilation. The percentage of Chicago area hospitals with priority groups was 86%, much higher than the 23% of US state ventilator allocation guidelines which recommend priority groups.^9^ One reason for this difference may be that individual hospitals feel a higher obligation to give priority to members of their own medical staff whom they are directly responsible for when compared to the creators of US state guidelines. Of the hospitals with ventilator allocation guidelines, 71% chose to prioritize healthcare workers. Some hospitals prioritized multiple groups including one hospital that gives priority to pregnant patients, health care workers, essential workers, and families of essential workers. With so many prioritized groups, there is a concern that anyone not in one of these priority groups may not have equitable access to mechanical ventilation if a shortage occurs.^25^

The second survey showed that there was little agreement among respondents for how hypothetical patients should be ranked for priority, and almost half of all respondents resorted to tiebreakers in order to rank the patients. It seems doubtful that a committee writing an allocation protocol intended for tiebreakers to be used almost 50% of the time. If operationalized, arguably these carefully thought-out, elaborate protocols would actually function more like a lottery.

Some have maintained that a lottery system would ultimately be less likely to promote inequities.^4^ However, others submit that a lottery system would result in more deaths and actually not mitigate existing disparities.^24^

Results from the second survey did reveal that respondents agreed most about giving the 35-year-old city bus driver who had stage IV lung cancer and a SOFA score of 12 low priority. This patient was ranked last over 70% of the time (**Figure 1**), showing that the concept of likelihood to benefit from the ventilator is considered one of the top factors by most allocation protocols.

These results have significant implications. If critical care resources in this region are scarce, patients might seek out hospitals where they would receive higher priority. This could create an imbalance in hospital usage that further exacerbates resource scarcity within a hospital or healthcare system. Perhaps this is unlikely given that lack of transparency in hospital allocation protocols, as this study found no allocation protocols publicly available. Nevertheless, if groups within communities share experiences and unveil these differences, this could lead to mistrust and concerns about inequitable treatment of certain populations. Given the existing mistrust of healthcare systems among certain communities, exacerbated by differences in disease burden and mortality among Black, Latinx, and Native American populations during the COVID-19 pandemic,^26,27^ avoiding further erosions in confidence is crucial. A potential solution is to bolster regional guidance for resource allocation protocols based on working groups with representation from local hospitals.

Limitations of the study include a low response rate for Survey 1, which may be related hospitals lacking ventilator allocation policies or reluctance from some hospitals to share information about their protocols for research analysis. Another limitation is that the survey data may not reflect the current resource allocation protocols as many hospitals are continually updating them as issues of health disparities and difficulties with applying the protocols are raised,^24^ and as knowledge about COVID-19 and treatments increases. As all hospitals surveyed were in the Chicagoland area, the variability in allocation methods may not be generalizable to other metropolitan areas where increased state guidance was provided to hospitals when allocation policies were developed.^9,28^ In addition, some hospital protocols may provide triage officers with additional patient data not provided in the descriptions of the hypothetical patients when determining patient priority. Triage officer variation could only be described within two hospitals due to the low number of respondents from other hospitals, which may be related to lack of establishing triage officers at some hospitals. Finally, in many hospitals, a triage officer is unlikely be the sole decision-maker when prioritizing patients. Rather, a triage group or committee may be working together to determine patient priority and therefore, discussion and elaboration of patient data may make their choices more internally consistent.

## Conclusion

Although most Chicago area hospitals surveyed created a ventilator allocation guideline for use during the COVID-19 pandemic, these guidelines varied significantly and would lead to non-uniform allocation of ventilators amongst hospitals. Furthermore, the application of the hospital protocols was extremely varied and led to different prioritization of hypothetical patients within and across hospitals. The potential impact of such variation is inequitable distribution of resources further exacerbating community mistrust and disparities in health. Efforts to minimize this variation are needed.

## Supporting information

Supplemental Content

## Data Availability

Data referred to in the manuscript is available within the manuscript and the supplemental content.

## Author Contribution

RG, GP, WP and KM contributed substantially to the conception, study design, data analysis and interpretation. RG and GP coauthored the initial draft of the manuscript. WP and KM contributed substantially to editing and revising the manuscript. All authors give final approval of the version to be published. RG is the guarantor of the content of the manuscript, including the data and analysis. The authors appreciate the contributions provided by: Elaine Morgan, Eric Swirsky, Erin Paquette, Lainie Ross, Clint Moore, and Lisa Anderson who aided in the conception of this project.

