## Supplemental Content for "Regional Variation in COVID-19 Scarce Resource Allocation Protocols"

e-Table 1: Hospital Characteristics

| | Public or Private | Safety Net Hospital | Academic or Community | Trains medical students, residents or fellows | Religious Affiliation | Clinical Trials Available for Patients with COVID-19 | Location of hospital or hospital system |
|---|---|---|---|---|---|---|---|
| Hospital 1 | Private | No | Community | Yes | Yes | No | Urban, Suburban |
| Hospital 2 | Public | Yes | Academic and Community | Yes | Yes | Yes | Urban |
| Hospital 3 | Private | Yes | Academic and Community | Yes | Yes | Yes | Urban, Suburban, Rural |
| Hospital 4 | Private | No | Academic | Yes | No | Yes | Urban, Suburban |
| Hospital 5 | Private | No | Academic | Yes | No | Yes | Urban |
| Hospital 6 | Private | No | Academic | Yes | Yes | Yes | Suburban |
| Hospital 7 | Private | No | Academic | Yes | No | Yes | Urban |
| Hospital 8 | Public | Yes | Academic | Yes | No | Yes | Urban |

e-Table 2: Adult Exclusion Criteria

| | |
|---|---|
| Hospital 1 | None |
| Hospital 2 | None |
| Hospital 3 | Any code status order as determined by the patient and treating physician that would prohibit treatment involving the scarce resource, Intractable hypotension despite maximum dosage of two or more vasopressors, Cardiac arrests, Persistent coma or vegetative state, Acute severe neurological event with minimal chance of functional neurological recovery, Severe burns with less than 10% chance of survival, |
| Hospital 4 | None |
| Hospital 5 | 1) pediatric priority over adults, 2) adults with irreversible/advantaged neurologic condition, unwitnessed cardiac arrest with poor neurologic prognosis per AAN guidelines, severe burn with <10% survival, cardiac arrest unresponsive to CPR, any other condition resulting in near immediate death even with maximally aggressive therapy |
| Hospital 6 | Recurrent cardiac arrest, Active DNI orders, severe burns where predicted survival <10%, aneurysmal subarachnoid hemorrhage Hunt Hess Grade 5, ICH with ICH score 5 or higher, major hemispheric ischemic stroke with secondary brainstem injury and not eligible for decompressive hemicraniectomy, any other conditions with predicted immediate or new-immediate mortality even with aggressive therapy |
| Hospital 7 | None |
| Hospital 8 | None |

e-Table 3: Priority Groups for Pregnancy, Health Care Workers, and Essential Worker

| | Pregnancy | How Priority Occurs (Pregnancy) | Health Care Workers | How Priority Occurs (Health Care Workers) | Essential Workers | How Priority Occurs (Essential Workers) |
|---|---|---|---|---|---|---|
| Hospital 1 | Yes | Automatically in highest priority category (age of fetus not mentioned) | No | N/A | No | N/A |
| Hospital 2 | N/A | N/A | N/A | N/A | N/A | N/A |
| Hospital 3 | Yes | If woman is greater than 26 weeks pregnant with viable fetus, she will have one point subtracted from her SOFA score | Yes | Priority in setting of a tie | No | N/A |
| Hospital 4 | Yes | Considered separate from main scoring system | Yes | Subtract one point from scoring system | Yes | Subtract one point from scoring system |
| Hospital 5 | Yes | Pregnant women given priority in setting of a tie | Yes | Priority in setting of a tie | Yes | Priority in setting of a tie |
| Hospital 6 | Yes | Pregnant women with viable fetus given priority in setting of a tie | Yes | Priority in setting of a tie | No | N/A |
| Hospital 7 | No | N/A | No | N/A | No | N/A |
| Hospital 8 | Yes | Not discussed | Yes | Not discussed | No | N/A |

e-Table 4: Priority Groups for Families of Essential Workers, Pediatric Patients

| | **Families of Essential Workers** | **How are families of essential workers given priority** | **Pediatric Patients** | **How are pediatric patients given priority** |
|---|---|---|---|---|
| Hospital 1 | No | N/A | No | N/A |
| Hospital 2 | No | N/A | No | N/A |
| Hospital 3 | No | N/A | No | N/A |
| Hospital 4 | Yes | Subtract one point from scoring system | No | N/A |
| Hospital 5 | No | N/A | Yes | Given priority over adult patients |
| Hospital 6 | No | N/A | No | N/A |
| Hospital 7 | No | N/A | No | N/A |
| Hospital 8 | No | N/A | No | N/A |

e-Table 5: Adult Withdrawal of Mechanical Ventilation

| | Protocol discusses withdrawing mechanical ventilation from one patient to another | Protocol for considering withdrawal of mechanical ventilation | Who leads discussion about withdrawal of mechanical ventilation with family | How to talk with family about withdrawal of mechanical ventilation discussed |
|---|---|---|---|---|
| Hospital 1 | Yes | Not discussed | Not discussed | Not discussed |
| Hospital 2 | N/A | N/A | N/A | N/A |
| Hospital 3 | Yes | Not discussed | Not discussed | Yes |
| Hospital 4 | Yes | Reallocation based on SOFA scores and additional assessment of survivability made by triage team and intensivist attending | Triage committee | Not discussed |
| Hospital 5 | Yes | Withdraw occurs if the new arriving patient has a SOFA score at least 3 points lower than a currently ventilated patient | Not discussed | Yes |
| Hospital 6 | Yes | Triage chair identifies a patient with SOFA >16, notified attending intensivist, decision made to withdraw mechanical ventilation, no appeals process | Triage chair | Not discussed |
| Hospital 7 | Yes | After 48 hours, if patient in lower priority category (yellow or orange), can have ventilator removed if patient in highest priority category (red) in need of a ventilator. Lower priority categories include patients with high priority score and no improvement in SOFA score in past 48 hours. | Primary attending for patient | Yes |
| Hospital 8 | No | No | No | No |

e-Table 6: Hospitals with Unique Criteria for Pediatric Patients

| | Age of Pediatric Patients | Exclusion Criteria | What are the pediatric exclusion criteria? | What scoring system is used to triage pediatric patients? | Is withdrawal of mechanical ventilation discussed for pediatric patients? | How is pediatric ventilator withdrawal explained to occur (i.e. what is the actual mechanism for withdrawal, how it is discussed with family)? | Are pediatric patients triaged with adults? |
|---|---|---|---|---|---|---|---|
| Hospital 3 | Less than 18 | Yes | NICU patients are excluded | pSOFA | Yes | Education provided via simulation on how to have this discussion. Talking points in the protocol. | No |
| Hospital 5 | Less than 18 | No | N/A | No scoring system, automatically get priority over adults | No | Not discussed | No |
| Hospital 6 | Less than 18 | Yes | Recurrent cardiac arrest, active DNI orders, severe burns where predicted survival <10%, aneurysmal subarachnoid hemorrhage Hunt Hess Grade 5, ICH with ICH score 5 or higher, major hemispheric ischemic stroke with secondary brainstem injury | Clinical judgment | No | Not discussed | Yes |

| | | | | | | | |
|---|---|---|---|---|---|---|---|
| | | | and not eligible for decompressive hemicraniectomy, any other conditions with predicted immediate or near-immediate mortality even with aggressive therapy | | | | |
| Hospital 7 | Not mentioned | No | N/A | Clinical judgment: | No | Not discussed | Yes |

pSOFA-pediatric sequential organ failure assessment

e-Table 7: Adult Tiebreakers:

| | 1st tiebreaker | 2nd tiebreaker | 3rd tiebreaker | 4th tiebreaker |
|---|---|---|---|---|
| Hospital 1 | Not discussed | Not discussed | Not discussed | Not discussed |
| Hospital 2 | N/A | N/A | N/A | N/A |
| Hospital 3 | Greatest likelihood of positive clinical outcome | Priority to healthcare/frontline workers | Random allocation | N/A |
| Hospital 4 | Interventional COVID-19 research subjects | Hospital system front line workers | Priority to younger patients | Random allocation |
| Hospital 5 | Essential workers | Pregnant women | Priority to younger patients | Random allocation |
| Hospital 6 | Lottery | N/A | N/A | N/A |
| Hospital 7 | Priority to younger patients | Raw priority score | Random allocation | N/A |
| Hospital 8 | Committee evaluation | N/A | N/A | N/A |

e-Table 8: Creation of the Ventilator Allocation Policy

| | **Who were involved in creation the protocol (i.e. hospital administrators, ethics, critical care physicians, chaplains...)?** | **Did you trial use of this protocol at your hospital?** | **Did you need to use the protocol at your hospital due to an internal ventilator shortage?** |
|---|---|---|---|
| Hospital 1 | Ethics, critical care physicians, nurses, hospital administrators, system director of diversity & inclusion | No | No |
| Hospital 2 | N/A | N/A | N/A |
| Hospital 3 | System resource allocation team physician leader, supply chain, ethics, critical care physician, nursing leader, emergency medicine physician, infectious disease physician, diversity and inclusion representative, legal, palliative care clinician, pharmacy, care management, spiritual care. | Yes | No |
| Hospital 4 | A 15 member system wide team of administration, physicians, nurses, chaplains, ethics | No | No |
| Hospital 5 | Broad committee including ethics, critical care, palliative care, infectious disease, hospital leadership | Yes | No |
| Hospital 6 | Physicians, ethicists, administrators, clinical leadership, critical care committee, and med exec committee, legal | No | No |
| Hospital 7 | Hospital administrators, ethics, critical care physicians, geriatric physician, legal | No | No |
| Hospital 8 | Representatives from critical care, emergency medicine, trauma, ethics | No | No |

e-Table 9: Community Involvement with Creation of Ventilator Allocation Policy

| | **Were community representatives involved in the formation of the protocol?** | **Which community members were involved?** | **How were community members involved?** | **How did your institution plan to explain the triage mechanism for mechanical ventilation to the public?** | **Are any of your institution's policies on allocation of scarce resources publicly available?** |
|---|---|---|---|---|---|
| Hospital 1 | No | N/A | N/A | Not discussed | No |
| Hospital 3 | No | N/A | N/A | Had explanatory document ready but not provided to public as ventilator shortage did not occur | No |
| Hospital 4 | No | N/A | N/A | No plan in place | No |
| Hospital 5 | Yes | An existing community advisory council created to advise the hospital | The community advisory council reviewed the protocol framework and provided critical feedback | No plan in place | No |
| Hospital 6 | No | N/A | N/A | Planned to discuss with individual admitted patients if shortage occurred | No |
| Hospital 7 | No | N/A | N/A | Had explanatory document for ready but not provided to public as ventilator shortage did not occur | No |
| Hospital 8 | Yes | Not discussed | Not discussed | Not discussed | No |

e-Table 10: Triage Committee Composition

| | Triage Committee Created | Critical Care Physician on Committee | Ethicist on Committee | Nurse on Committee | Others Included on Committee |
|---|---|---|---|---|---|
| Hospital 1 | Yes | Not discussed | Not discussed | Not discussed | Not discussed |
| Hospital 2 | No | N/A | N/A | N/A | N/A |
| Hospital 3 | Yes | Yes | Yes | Yes | system resource allocation team physician, supply chain, emergency medicine physician, infectious disease physician, diversity and inclusion representative, legal, palliative care clinician, pharmacy, care management, spiritual care, chief medical officer/delegate, director of nursing/delegate, respiratory therapy lead/delegate, clinical risk |
| Hospital 4 | Yes | Yes | Yes | No | Non-intensivist physician, hospital representative |
| Hospital 5 | Yes | Yes | No | No | N/A |
| Hospital 6 | Yes | Yes | Yes | Yes | N/A |
| Hospital 7 | Yes | Yes | Yes | Yes | infectious disease physician, hospital administration representative |
| Hospital 8 | No | N/A | N/A | N/A | N/A |

e-Table 11: Triage Committee Guidelines

| | Who is restricted from being on triage committee | Information provided to triage committee | Information blinded from triage committee | How is information provided to triage committee |
|---|---|---|---|---|
| Hospital 1 | Not discussed | MSOFA score, major comorbidities | Not discussed | Primary medical team |
| Hospital 2 | N/A | N/A | N/A | N/A |
| Hospital 3 | Bedside clinician | MRN, SOFA score, pregnancy status, and severe comorbid conditions with death likely within one year | Yes (blinded information not identified) | Risk managers important information to a spreadsheet. |
| Hospital 4 | Bedside clinician | SOFA score and other information that is used to calculate priority score. Discussion with primary intensivist attending if reallocation of ventilators needed. | Not discussed | Spreadsheet generated that triage team can review chart and contact primary medical team for necessary data |
| Hospital 5 | Bedside clinician | Full access to the electronic medical record | No | Triage committee has full access to medical record |
| Hospital 6 | Not discussed | Age, SOFA score, and priority conditions (pregnancy and healthcare worker status) | Yes (patient name, bed location, medical service, race, ethnicity) | List in medical record |
| Hospital 7 | Bedside clinician | Priority score, intubation status | Not discussed | Primary team to update priority score for their patients in the medical record which is provided to triage committee |
| Hospital 8 | N/A | N/A | N/A | N/A |

e-Table 12: Other Scarce Resource Protocols

| | **Did your institution create protocols for allocation of scarce resources other than ventilators (i.e. Remdesivir, ECMO, blood products, dialysate solution)?** | **If yes, what protocols were created?** | **Which, if any, of these protocols were used in practice?** |
|---|---|---|---|
| Hospital 1 | Yes | Remdesivir, protocols for allocation of other scarce drugs (not identified) | Not discussed |
| Hospital 2 | No | N/A | N/A |
| Hospital 3 | Yes | Remdesivir, ECMO, CRRT, Blood products, HiFlow/Bipap, testing | Remdesivir |
| Hospital 4 | Yes | Remdesivir | Remdesivir |
| Hospital 5 | Yes | Remdesivir, ECMO | ECMO |
| Hospital 6 | No | | Not discussed |
| Hospital 7 | Yes | Remdesivir, ECMO | ECMO |
| Hospital 8 | Yes | Dialysis | Not discussed |

*Aim 1 Survey: COVID-19 Related Protocols*

Ventilator Allocation Protocols

1. Did your institution create a ventilator allocation protocol?

Yes
No

2. If your institution did not have a ventilator allocation protocol, how was ventilator allocation planned to occur?

3 Are there exclusion criteria? (Exclusion criteria are criteria designed to remove patients from consideration who have a very low chance to benefit from mechanical ventilation. Exclusion criteria can also include groups that are given a ventilator regardless of priority score, such as children at some hospitals. These exclusion criteria occur first prior to scoring patients for a ventilator.

Yes
No

4. What are the exclusion criteria?

5. What initial scoring system is used (i.e. SOFA score, SOFA score plus evaluation of medical comorbidities)?

6. Does the protocol give priority to certain groups (i.e. healthcare workers, pregnant patients)?

Yes
No

7. If yes, what are these groups and how are they given priority?

8. How does the protocol handle tiebreakers?

9. Does the protocol involve the creation of a triage committee?

Yes
No

10. Are there any restrictions in who these triage committee members can be? (Restriction examples may include that triage committee members cannot care for patients at bedside, must be from certain specialties (i.e. critical care)...

11. Are triage committee members blinded to certain patient data (i.e. patient race, age)?

12. What information about patients is provided to committee members?

13. How is information about patient data provided to triage officers (i.e. by primary medical team, by IT uploaded of data from the medical record)?

14. Does the protocol discuss withdrawal of mechanical ventilation from one patient to provide to another at higher priority?

Yes
No

15. How is withdrawal of mechanical ventilation explained to occur (i.e. what is the actual mechanism for withdrawal, how it is discussed with family)?

Pediatric Ventilator Allocation Protocol

16. Does the protocol have unique criteria for pediatric patients?

Yes
No

17. What age is considered pediatric?

18. Do exclusion criteria exist for pediatric patients?

Yes
No

19. What are the pediatric exclusion criteria?

20. What scoring system is used to triage pediatric patients?

21. Is withdrawal of mechanical ventilation discussed for pediatric patients?

Yes
No

22. How is pediatric ventilator withdrawal explained to occur (i.e. what is the actual mechanism for withdrawal, how it is discussed with family)?

23. Are pediatric patients triaged with adults?

Yes
No
Creation and Use of Ventilator Allocation Protocol

24. Were community representatives involved in the formation of the protocol?

Yes
No

25. Which community members were involved?

26. How were community members involved?

27. How did your institution plan to explain the triage mechanism for mechanical ventilation to the public?

28. Are any of your institution's policies on allocation of scarce resources (i.e ventilator allocation, Remdesivir allocation...) publicly available?

Yes
No

29. Which policies are publicly available and by what means are they available?

30. Who were involved in creation the protocol (i.e. hospital administrators, ethics, critical care physicians, chaplains...)?

31. Did you trial use of this protocol at your hospital?

Yes
No

32. Did you need to use the protocol at your hospital due to an internal ventilator shortage?

Yes
No

Other Scarce Resource Protocols

33. Did your institution create protocols for allocation of scarce resources other than ventilators (i.e. No Remdesivir, ECMO, blood products, dialysate solution)?

Yes
No

34. If yes, what protocols were created?

35. Which, if any, of these protocols were used in practice?

36. If your institution does not have other scarce resource allocation protocols, how was/is allocation of these resources to occur?

Unilateral DNR Policy

37. Does your institution have a unilateral DNR policy?

Yes
No

38. Was your institution's unilateral DNR policy created or updated during the COVID-19 pandemic?

Yes
No

39. Does the policy limit unilateral DNR use to certain patients (i.e. patients with COVID-19, patients with an expected cardiac arrest with no reversible etiology)?

Yes
No

40. Is this policy active at all times, or only during the COVID-19 pandemic?

Active at all times
Only active during the COVID-19 pandemic

41. Which patients does the policy limit use of unilateral DNRs?

42. Does the policy require oversight for use of unilateral DNRs (such as two physicians to agree to the use of the unilateral DNR or require ethics committee approval)?

Yes
No

43. If yes, please explain the oversight required (such as if 2 physicians or ethics committee approval is needed).

44. Does the policy require documentation in the medical record about the use of the unilateral DNR?

Yes
No

45. Does your institution require placement of a DNR order or a unilateral DNR order?

Yes
No

46. Please describe what order is used in the medical record (i.e. a regular DNR order, or unilateral DNR order).

47. Does the policy require the physician discuss or attempt to discuss use of the unilateral DNR with the patient or alternate decision maker?

Yes
No

48. Is your unilateral DNR policy publicly available?

Yes
No

49. How is the policy public? (i.e. is it available on your institution's website, is it provided to patients and families on admission...)

50 Does the policy have other components not listed in the above questions? If yes, please explain.

51. Characteristics of your Institution (please check all that apply)

Public
Private
Safety net
Community
Academic
Trains medical students, residents, or fellows
Has a religious affiliation
Clinical trials for COVID-19 available to patients?
Urban
Suburban
Rural

52. In which COUNTY in Illinois is your hospital located?

Demographics of Person Completing Survey

53. Role at Hospital

*Aim 2 Survey: Triage Officer Allocation of Mechanical Ventilation during the COVID-19 Pandemic*

1. Were you notified by your hospital that you would be a triage officer or on a hospital resource allocation team to decide how to prioritize scarce resources?

Yes
No

2. To your knowledge, does your institution have a written protocol that describes how to allocate scarce resources such as mechanical ventilators?

Yes
No
Unsure

3. How familiar are you with this resource allocation document prior to receiving this survey?

Very familiar (received extensive training or helped create it
Somewhat familiar (received some training)
Neutral (I know where to find the document or who to ask if needed)
Not familiar

4. Below is a list of 6 hypothetical patients. Please read each one and rank them according to the priority these patients would receive under your hospital's resource allocation document.

All 6 patients arrived within 5 minutes of each other, with patient 1 being the first to arrive followed by patient 2, 3, 4, 5, and 6 in that order.

Please take into account anything that might be mentioned in your hospital's resource allocation document such as: exclusion criteria, priority scoring system used (i.e.SOFA), chronic conditions, age, pregnancy, reciprocity etc.

If one or more of the patients listed below would have been excluded from priority scoring at your institution, please rank them anyway. (Examples of patients meeting exclusion criteria may include patients who would receive a ventilator without needing a priority score such as pediatric patients at some hospitals, or patients who would not receive a ventilator regardless of priority score such as patients age >90 at some hospitals)

If you needed to use a random lottery as a tiebreaker, please complete the tiebreaker using a random number generator (such as https://bit.ly/2V1jKXm).

Assume that all SOFA scores were calculated at the same time interval.

Please rank all 6 patients below from 1 (highest priority for a ventilator ) to 6 (lowest priority for a ventilator). Do the best that you can ranking these patients with the information provided.

A. 91-year-old retired doctor with a past medical history of coronary artery disease, history of stroke 10 years ago with no residual disease. SOFA: 6

B. 65-year-old retired day-care teacher with a past medical history of poorly controlled diabetes. SOFA: 7

C. 45-year-old ED nurse with active breast cancer (receiving chemotherapy with curative intent) and is currently immunocompromised. SOFA: 11

D. 35-year-old city bus driver with stage IV lung cancer, participating in clinical trial for lung cancer through the medical center where he receives care. SOFA: 12

E. 25-year-old graduate student with hypertension, 30 weeks pregnant. SOFA: 14

F. 12-year-old with a past medical history of hypoxic ischemic encephalopathy resulting in severe cognitive disabilities and severe asthma. She does not speak, is wheelchair bound, and communicates with her family by smiling. Her mother is a physician. SOFA: 3, PELOD-2: 8, pSOFA: 5, estimated mortality < 5%

5. Do any of these patients meet exclusion criteria in your institution's protocol?

Examples of patients meeting exclusion criteria could include patients receiving a ventilator without needing a priority score (i.e. pediatric patients as some hospitals), or patients not receiving a ventilator regardless of priority score (i.e. patients age >90 at some hospitals)

Yes
No

6. Which patients met exclusion criteria and what criteria did they meet?

7. Did you use any tiebreakers when ranking the above patients?

Yes
No

8. Which tiebreakers did you need to use and which patients did you use it on?

9. Do you believe your institutions policy provides sufficient guidance to rank the above patients?

Yes
No

10. If no, what in your institution's guidelines needs to be added or changed to make the guideline sufficient to rank the above patients?

11. Do you believe your institution's policy disadvantages any group (such as racial minorities, elderly age…)?

Yes
No

12. If yes, which groups are disadvantaged in your opinion?

Racial/ethnic minority groups
People from low socioeconomic status
Elderly age
People with comorbidities
People with disabilities
Other

13. If other, please explain which groups you believe are disadvantaged

14. Are you in agreement with groups given priority for mechanical ventilation at your institution (i.e. healthcare workers, pregnant patients, pediatric patients)?

Yes
No
Our institution does not give priority to any group

15. If not, please explain which priority groups you do not agree with.

16. What is your profession (please select all that apply)?

Nurse
Physician
Hospital Administrator
Ethics
Other

17. If other, please describe your profession
